# The bidirectional causal effects of brain morphology across the life course and risk of Alzheimer’s disease: A cross-cohort comparison and Mendelian randomization meta-analysis

**DOI:** 10.1101/2021.05.14.21256707

**Authors:** Roxanna Korologou-Linden, Bing Xu, Elizabeth Coulthard, Esther Walton, Alfie Wearn, Gibran Hemani, Tonya White, Charlotte Cecil, Tamsin Sharp, Henning Tiemeier, Tobias Banaschewski, Arun L.W. Bokde, Erin Burke Quinlan, Sylvane Desrivières, Herta Flor, Antoine Grigis, Hugh Garavan, Penny Gowland, Andreas Heinz, Rüdiger Brühl, Jean-Luc Martinot, Marie-Laure Paillère Martinot, Eric Artiges, Frauke Nees, Dimitri Papadopoulos Orfanos, Tomáš Paus, Luise Poustka, Sabina Millenet, Juliane H. Fröhner, M Smolka, Henrik Walter, Robert Whelan, Gunter Schumann, Laura D Howe, Yoav Ben-Shlomo, Neil M Davies, Emma L Anderson

## Abstract

Neuropathological changes associated with Alzheimer’s disease (AD) can occur decades before clinical symptoms. We investigated whether neurodevelopment and/or neurodegeneration affects the risk of AD, through reducing structural brain reserve and/or accelerating brain atrophy, respectively. We used bidirectional two-sample Mendelian randomization to estimate the effects of genetic liability to AD on global and regional cortical thickness, total intracranial volume, volume of subcortical structures and cerebral white matter in 36,842 participants aged eight to 81 years across five independent cohorts, and the effects of global and regional cortical thickness and subcortical volumes on AD risk in 94,337 participants. Our findings show that AD risk alleles have an age-dependent effect on a range of cortical and subcortical brain measures that starts in mid-life, in non-clinical populations. Evidence for such effects across childhood and young adulthood is weak. We also found little evidence to suggest brain morphology alters AD risk. Thus, genetic liability to AD is likely to alter mechanisms and/or rates of neurodegeneration, rather than reduce structural brain reserve.

## Main

The earliest Alzheimer’s disease (AD)-related histopathological changes are typically observed within the medial temporal lobes (e.g. entorhinal cortex, hippocampus) and disperse throughout the frontal, parietal and temporal neocortices and subcortical regions by the time a clinical diagnosis of AD is made^1^. Research into autosomal dominant forms of AD has shown that amyloid-β accumulation in the brain may be apparent 20 years before the appearance of clinical symptoms^2^. Consequently, the earliest evidence of AD pathology is not well captured using clinical symptoms. Integration of biological data prior to the onset of clinical symptoms is a crucial step in understanding the aetiology, timing, and progression of the disease, and for the development of more efficient strategies for early detection and screening of individuals for AD risk.

It has been argued that AD risk may be mediated through both morphology (“brain reserve”) and/or functional capacity to compensate for pathology (“cognitive reserve”)^3^,which may operate synergistically. Changes in brain structure may mediate the effect of such genetic variants on AD risk, through determining the underlying brain reserve of an individual. Furthermore, the relationship between brain structures and AD may be bidirectional, as genes associated with changes in brain morphology, such as thickness and surface area, have been shown to be involved in neurodevelopmental processes such as neurogenesis^4^. Genetic instruments allow for the identification of factors that modify disease risk, establish downstream effects of prodromal disease and discover biomarkers that predict disease. Genome-wide association studies (GWAS)^5–8^ for AD have identified approximately 30 single nucleotide polymorphisms (SNPs), each with a modest effect on the risk of AD, apart from the ε4 genotype in the *APOE* gene, whereby homozygous carriers have up to twelve-fold increased risk^8^. The heritability of AD is large, with estimates as high as 79%^9^, in comparison heritability of brain structures, such as average cortical thickness, has been estimated to be 25%^10^.

Mormino et al^11^ reported that an AD polygenic risk score (PRS) at p<0.01 for single nucleotide polymorphisms (SNP) inclusion was associated with lower hippocampal volume in cognitively unaffected older participants (age: 73.5-75.3 years) and participants with AD, but found little evidence that the PRS was associated with hippocampal volume in young adults (age: 18-35 years, N=1,322). However, these studies have limited sample sizes (between 104 and 1,024 participants) because genetic and neuroimaging data are rarely available in combination. The largest brain imaging GWAS to date^12^ used brain magnetic resonance imaging (MRI) scans from approximately 40,000 healthy individuals, combining study samples from the Enhancing Neuroimaging Genetics through Meta-Analysis (ENIGMA), the Cohorts for Heart and Aging Research in Genomic Epidemiology (CHARGE), and the first release of UK Biobank (UKB) imaging data. SNPs associated with brain structure have been discovered using far larger sample sizes than in previous neuroimaging studies, allowing for the investigation of the causal effects of structural brain measures on risk of AD, using Mendelian randomization (MR).

MR is a form of instrumental variable analysis which uses SNPs as instruments for environmental exposures^13^. MR can provide evidence of lifetime effects of phenotypes on disease risk (and vice versa) and, under various assumptions, is robust to many forms of bias prevalent in other observational study designs such as confounding, reverse causation and measurement error.

In our study, we investigated how genetic liability to AD affects brain morphology across the life course (from ages 8-81 years) using two-sample MR. This approach helps to test whether AD genetic susceptibility affects brain development or degeneration. Using two-sample MR, we also investigated whether brain morphology has a causal effect on the risk of AD, to establish whether greater thickness/volume provides a protective effect against advancing neuropathology and thus, reduces risk of an AD diagnosis (“brain reserve” hypothesis). A better understanding of the mechanisms through which genes are acting on AD, and the timing of this, may aid in the development of effective intervention strategies.

## Results

We used bidirectional two sample-MR^14^ to firstly examine the effect of SNPs that are robustly and independently associated with AD (p≤5×10^−8^) on global and regional cortical thickness, estimated total intracranial volume, and volumes of subcortical structures. We also included total white matter as an outcome where available. Measures for regional cortical thickness and subcortical structures were adjusted for mean thickness and total estimated intracranial volume, respectively. To boost the statistical power of the smaller childhood cohorts, we meta-analysed the causal effect estimates across ABCD^15,16^, Generation R^17,18^, and IMAGEN^19^ (ages 8-16 years). For early adulthood, we used participants selected for neuroimaging in ALSPAC sub-studies^20^ (age 18 to 24.5 years). For mid- to late-adulthood, we stratified the UK Biobank population into three age tertiles; 45-60 years, 60-68 years, and 68 to 81 years (Supplementary Methods). In total, we used 23-25 independent AD SNPs from the largest GWAS of clinically diagnosed AD^21^, depending how many were available in each cohort used (see Table 1 and Supplementary Data Tables 1-5). We have grouped the brain regions in order of Braak staging of tau pathology, approximating the anatomical definitions of transentorhinal (Braak stage I/II), limbic (III/IV), and isocortical (VI) Braak stages^22^. Braak staging is a system characterised by post-mortem autopsies by Braak and Braak and is based on the premise that AD pathological tau severity progresses in stages across locations, with severity of symptoms being correlated to the progression through the Braak stages^22,23^. Regions not included in the Braak staging have been grouped together at the end for completion.

**Table 1.** Descriptive statistics of the cohorts used in the analysis

| Cohort | N | No. of SNPs for Alzheimer's disease instrument | F-statistic for Alzheimer's disease instrument | Mean age (years) (SD) | Age range (years) | %female |
| --- | --- | --- | --- | --- | --- | --- |
| ABCD | 5,022 | 25 | 223.28 | 9.91 (0.62) | 8.92-11 | 52.6% |
| ALSPAC | 446-776 | 25 | 231.70 | 20.38 (1.45) | 18-24.5 | 27.7% |
| Generation R | 1,175 | 23 | 239.34 | 10.16 (0.61) | 8.71-11.99 | 49.2% |
| IMAGEN | 1,698-1,739 | 25 | 224.67 | 14.42 (0.39) | 12.94-16.04 | 50.4% |
| UK Biobank (entire) | 28,130 | 24 | 231.46 | 63.81 (7.50) | 45-81 | 52.2% |
| UK Biobank Tertile 1 | 9,377 | 24 | 231.46 | 55.09 (3.42) | 45-60 | 57.0% |
| UK Biobank Tertile 2 | 9,377 | 24 | 231.46 | 64.34 (2.24) | 60-68 | 53.7% |
| UK Biobank Tertile 3 | 9,376 | 24 | 231.46 | 72.01 (2.88) | 68-81 | 46.0% |

Secondly, we examined the causal effects of brain morphology on AD risk, using genetic instruments for cortical thickness and subcortical structures from the ENIGMA consortium GWAS (Supplementary Data Table 7), which controlled for mean thickness and estimated total intracranial volume. A summary of our study design is presented in Figure 1.

**Figure 1:**
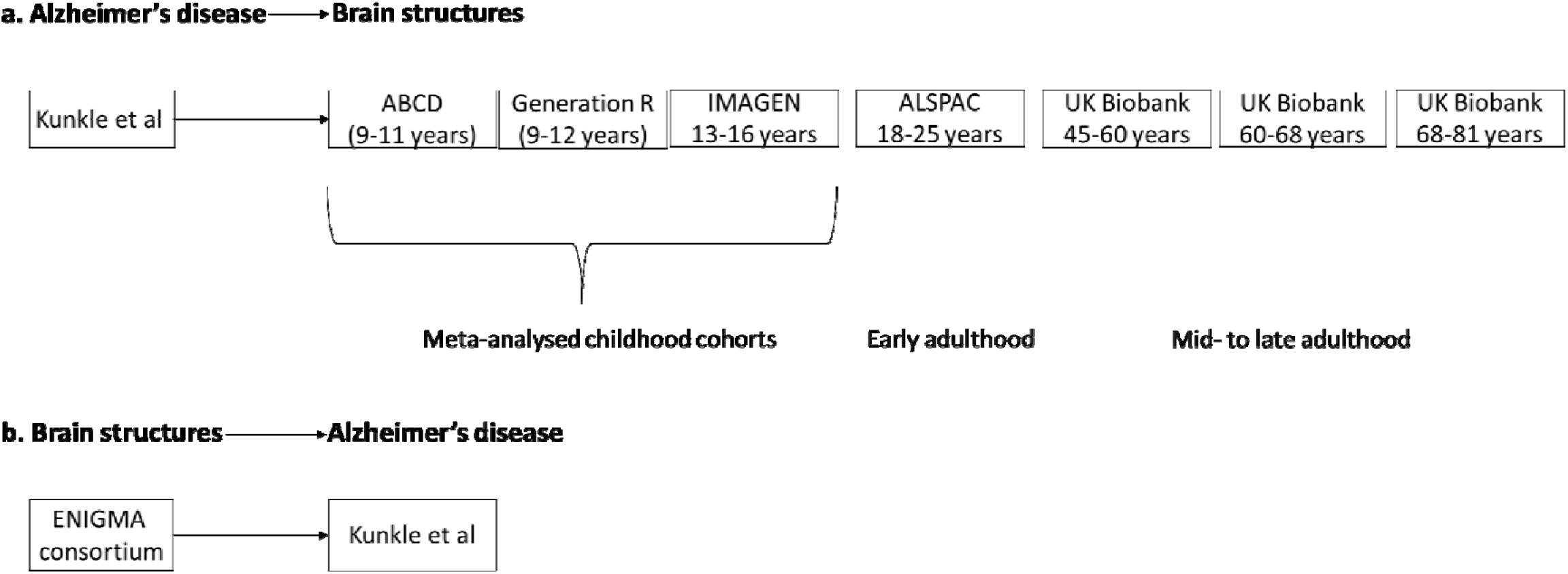
Diagram (a) describes our study design when conducting a Mendelian randomization study of AD on brain morphology. For childhood, we combined the effects estimated with the inverse variance weighted method for ABCD, Generation R, and IMAGEN, using a random effects model. Diagram (b) describes our study design when using Mendelian randomization of brain structures on AD, using summary-level data. In diagrams a and b, we test the hypotheses provided that the conditions (i), (ii), and (iii) are satisfied; the genetic proxy for an exposure is a valid instrument, in that (i) the SNPs for an exposure are strongly associated with the exposure they proxy (relevance), (ii) there are no confounders of the SNPs-outcome relationship (independence), and (iii) the SNPs only affect the outcome via their effects on exposure (exclusion restriction).

Of the 34 cortical regions and 10 subcortical structures examined, there was evidence to suggest that genetic liability to AD has an age-dependent effect on the thickness and volume of these measures respectively, across mid- to late adulthood, but the evidence for such effects in childhood through young adulthood is weak. The results for each age period are described in detail below. When we examined the causal effects of 22 cortical regions (i.e., those regions with genetic variants at 5×10^−8^), we found very little evidence of an effect of greater thickness on risk of AD. We only found evidence that hippocampal volume and thickness of lateral orbitofrontal and rostral anterior cingulate cortices affected the risk for AD.

### Causal effects of genetic liability to AD on brain structures

#### Childhood

Only weak evidence supported the association between genetic liability to AD and cortical thickness or subcortical volumes in school-aged children. A doubling in odds of genetic liability to AD was associated with a −0.01 standard deviation (SD) (95% confidence interval (CI): −0.02, −0.01) smaller volume of the hippocampus (Braak stage II) (Figure 2a), and −0.03 SD (95% CI: −0.05, −0.01) lower thickness of the caudal anterior cingulate (Braak stage IV) (Figure 2a, Supplementary Tables 1-4).

**Figure 2a.**
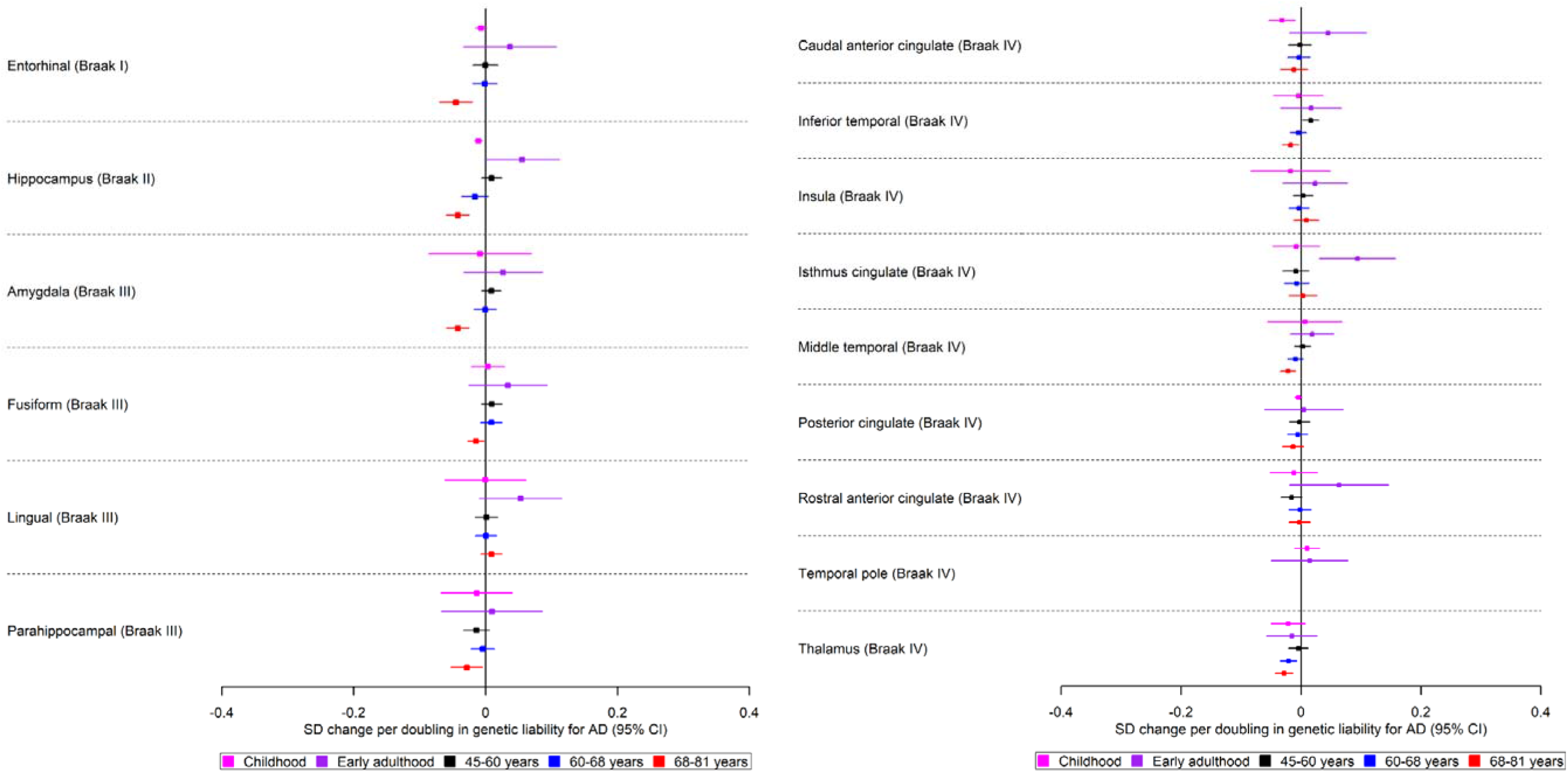
The causal effects of genetic liability to AD on brain structures in Braak stages I - IV at different ages across the life course (see Figure 2b for structures in Braak stage V and Figure 2c for Braak stage VI). The childhood cohorts include meta-analysed effects of three peri-pubertal cohorts: ABCD, GEN R and IMAGEN. The early adulthood cohort includes ALSPAC and the later adulthood cohorts include UK Biobank. Effect estimates for cortical regions and subcortical structures represent SD changes in thickness and volume. Cortical regions were adjusted for mean thickness and subcortical volumes were adjusted for estimated intracranial volume. Where an effect estimate is missing, that structural measure was not available in that cohorts.

#### Early adulthood

There was some evidence to suggest that higher genetic liability to AD is associated with regions affected in Braak stages IV and V (Figures 2a–2b). A doubling in odds of genetic liability to AD was associated with 0.05 SD (95% CI: −0.12, −0.01) lower thickness in the superior parietal (Figure 2a) and a 0.09 SD (95% CI: 0.04, 0.22) greater thickness in the isthmus cingulate cortex (Figure 2a, Supplementary Table 5).

**Figure 2b.**
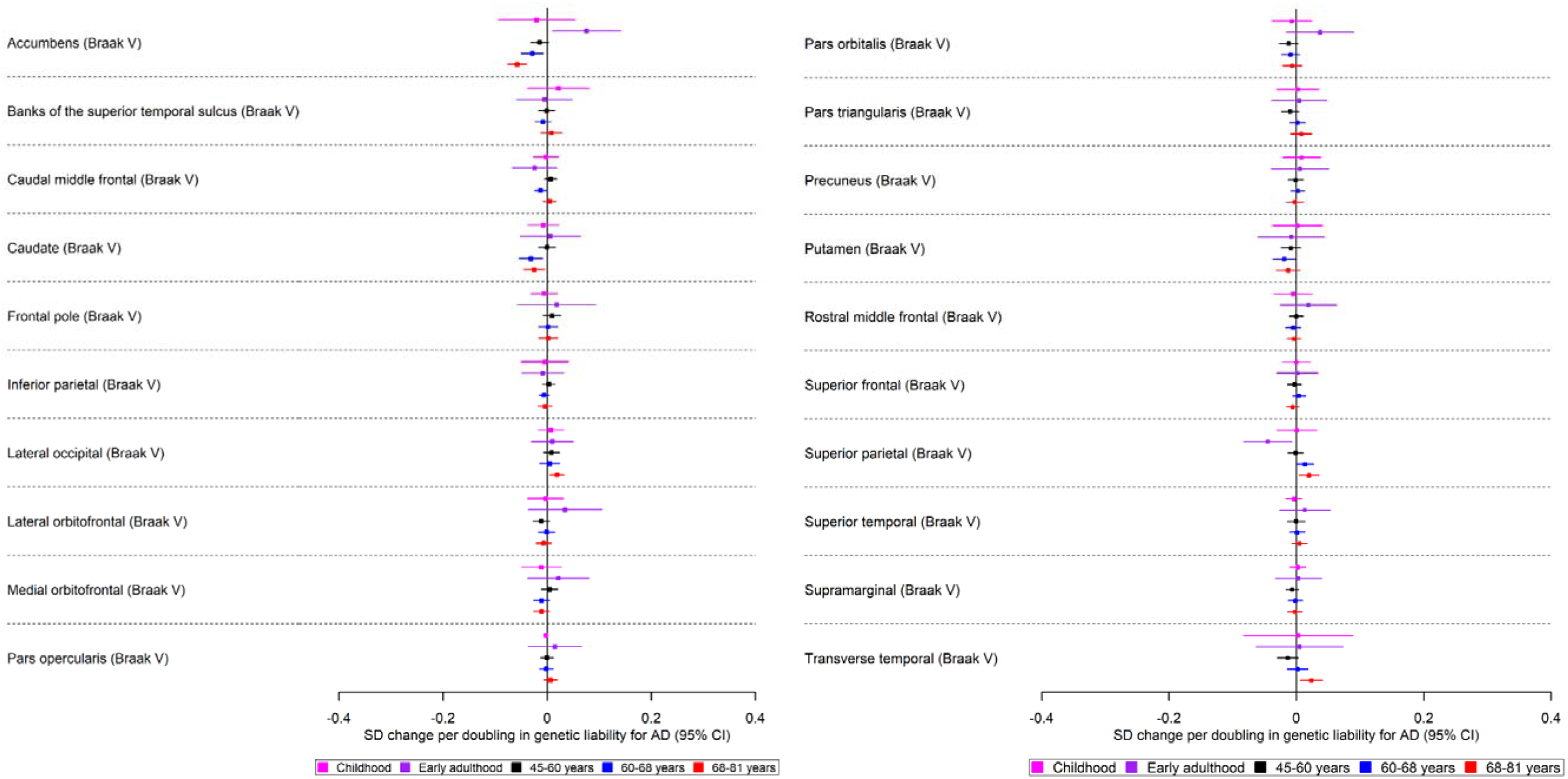
The causal effects of genetic liability to AD on brain structures in Braak stages V at different ages across the life course (see Figure 2c for structures in Braak stage VI). The childhood cohorts include meta-analysed effects of three peri-pubertal cohorts: ABCD, GEN R and IMAGEN. The early adulthood cohort includes ALSPAC and the later adulthood cohorts include UK Biobank. Effect estimates for cortical regions and subcortical structures represent SD changes in thickness and volume. Cortical regions were adjusted for mean thickness and subcortical volumes were adjusted for estimated intracranial volume. Where an effect estimate is missing, that structural measure was not available in that cohorts.

**Figure 2c.**
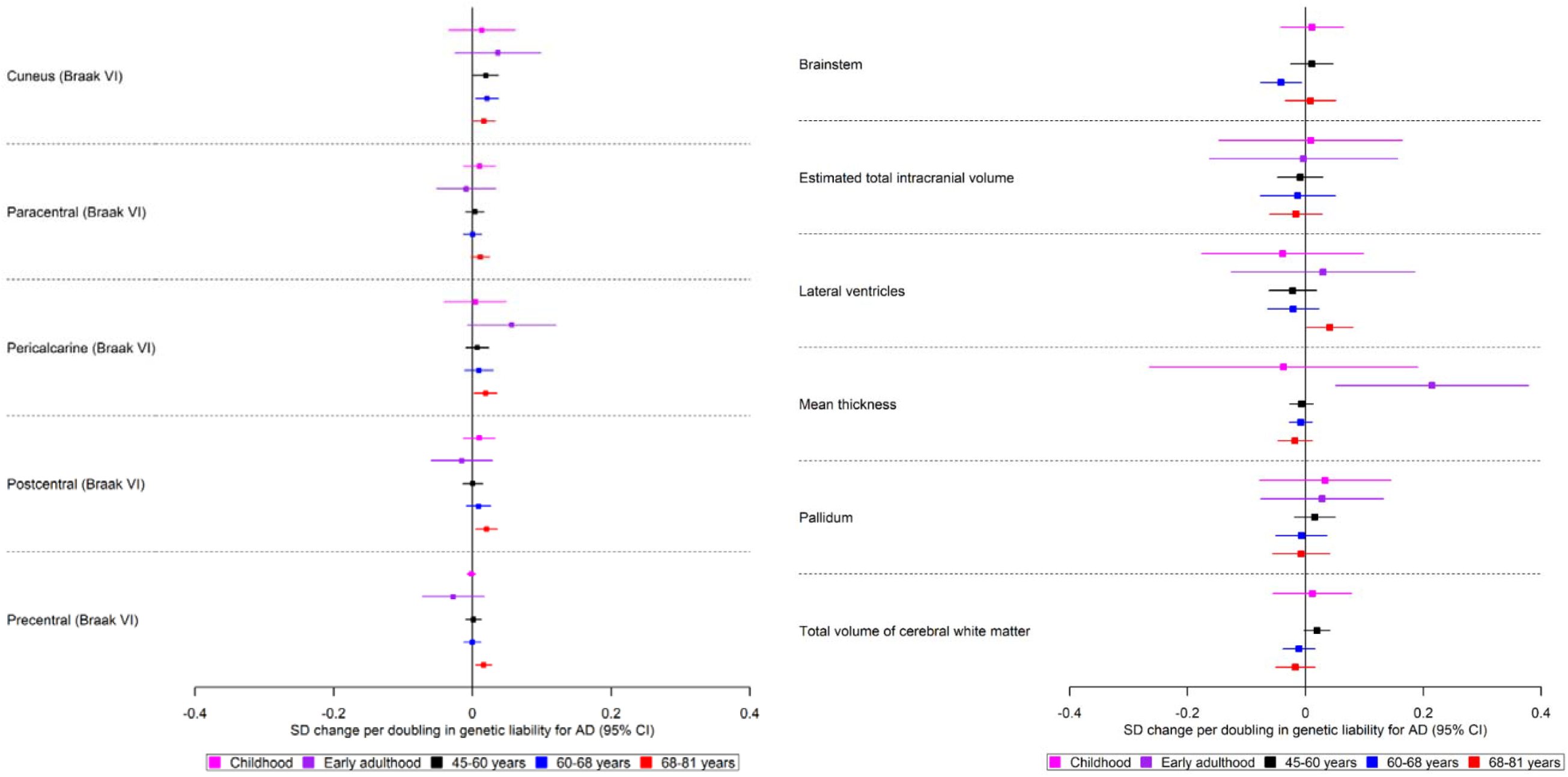
The causal effects of genetic liability to AD on brain structures in Braak stage VI, and those not included in Braak staging, at different ages across the life course. The childhood cohorts include meta-analysed effects of three peri-pubertal cohorts: ABCD, GEN R and IMAGEN. The early adulthood cohort includes ALSPAC and the later adulthood cohorts include UK Biobank. Effect estimates for cortical regions and subcortical structures represent SD changes in thickness and volume. Cortical regions were adjusted for mean thickness, subcortical structures and volume of cerebral white matter were adjusted for estimated intracranial volume. Where an effect estimate is missing, that structural measure was not available in that cohort.

#### Mid- to late life

We identified evidence of an age-dependent smaller volume of the hippocampus (Braak stage II), accumbens (Braak stage II), amygdala (Braak stage II), and thalamus (Braak stage IV) (p between age-stratified tertiles for each respective structure: 1.32×10^−5^, 0.001, 0.02 and 0.03; Figure 2a and Supplementary Tables 6 and 7). Furthermore, we found evidence of age-dependent lower thickness of the inferior temporal and middle temporal cortices (p between age tertiles=0.001 and p=0.009, respectively; Braak stage IV, Figure 2a). A higher genetic liability to AD, for example, was associated with 0.02 SD (95% CI: −0.04, −0.01) lower thickness in the middle temporal cortex for participants of aged 68-81 years and a trend in the same direction was observed for participants of age 60-68 years. On the contrary, for the superior and transverse temporal cortices (Braak stage V, Figure 2b), we identified a genetic liability to be associated with age-dependent greater thickness (p between age-stratified tertiles= 0.03 and p=0.003, respectively).

We also identified effects which did not show clear age-dependent associations. Within the youngest UK Biobank participants aged 45-60 years, a higher genetic liability to AD was associated with a greater thickness in the cuneus.

In participants aged 60 to 68 years, a higher genetic liability to AD was associated with a lower volume in the caudate (Braak stage V, Figure 2b). A higher genetic liability to AD was also associated with a smaller putamen volume, only in participants of this age group (Braak stage V, Figure 2b).

In participants aged 68-81 years, a doubling in odds of genetic liability to AD was associated with 0.05 SD (95% CI: 0.07, 0.02) lower thickness in the entorhinal cortex (Braak stage I). Additionally, a higher genetic liability to AD was associated with a lower thickness in the fusiform and 0.02 SD (95% CI: −0.03, −0.002) and parahippocampal cortices (Braak stage III, Figure 2a). A higher genetic liability was associated with a thicker pericalcarine, postcentral, precentral cortex and a larger volume in the lateral ventricles (Braak stage VI, Figure 2c).

#### Causal effects of brain morphology on risk of AD

We found little evidence of causal effects for the global measures of thickness and intracranial volume on AD risk (Supplementary Table 8). However, of the eight subcortical structures examined, we observed that a one SD increase hippocampal volume, instrumented by six SNPs, increased the risk for AD on average by 33% (95% CI:1.11,1.59). We also observed that a 1 SD increase in the thickness of the lateral orbitofrontal and rostral anterior cingulate cortices also increased risk of AD (OR: 2.74; 95% CI: 1.08, 6.93 and OR: 0.40; 95% CI: 0.19, 0.83, respectively, Figure 3). However, for these two structures, we have only one instrument, meaning we were unable to perform sensitivity analyses for assessing heterogeneity or pleiotropy and thus these results should be interpreted with caution.

**Figure 3.**
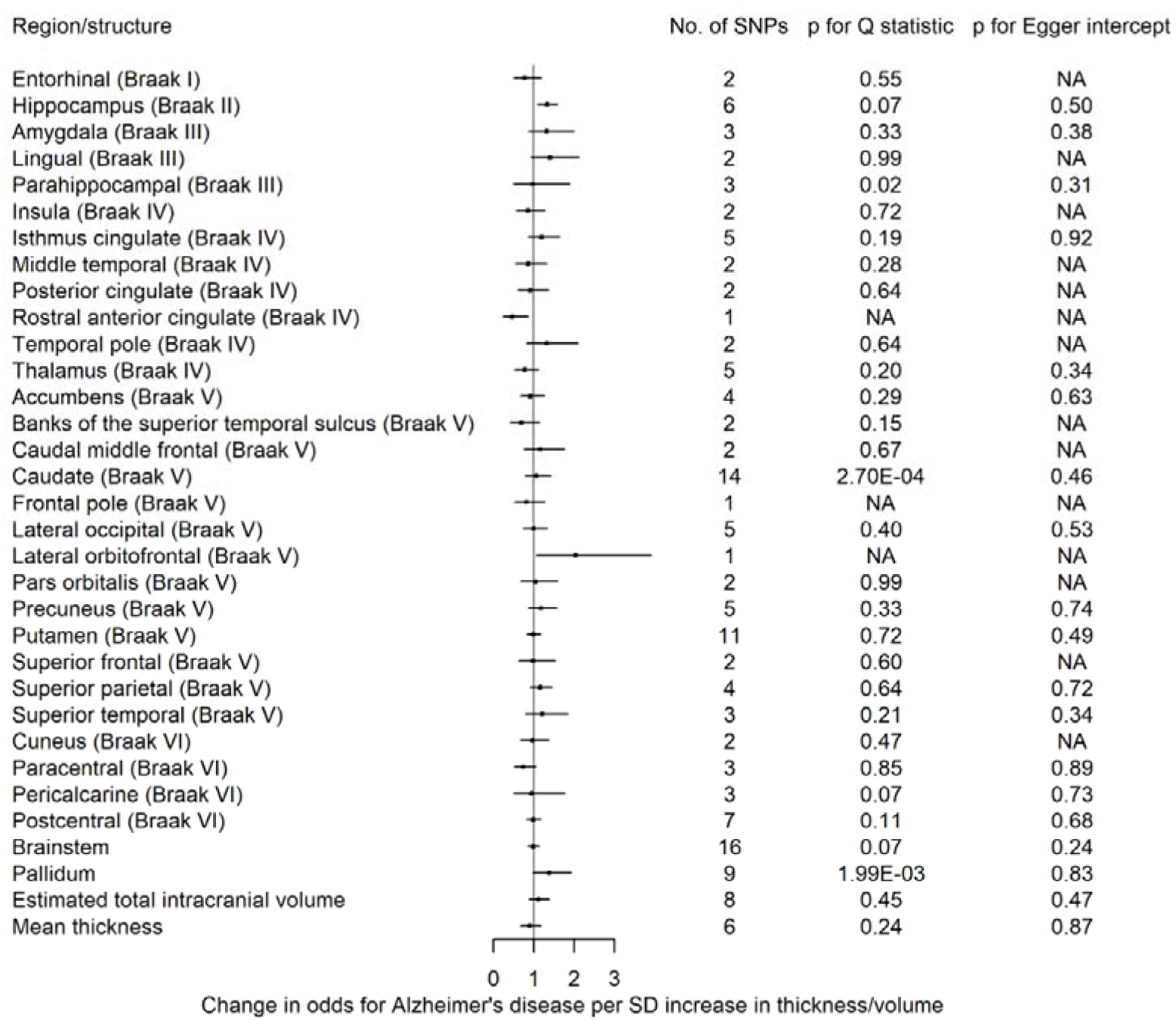
The causal effects of genetic predisposition to higher thickness and volume of cortical, subcortical and white matter measures, respectively on risk for AD. This figure shows the change in odds ratio for AD per standard deviation change in thickness and volume of cortical, subcortical structures, respectively. Effects for lateral ventricles is missing due to inability in obtaining access to summary statistics. The F-statistic is a measure of instrument strength.

#### Sensitivity analyses

Detailed results of sensitivity analyses measuring potential pleiotropy and instrument validity are in Supplementary tables 1-14. The evidence of a causal effect of genetic liability to AD on the caudal anterior cingulate in peri-pubertal childhood was consistent across pleiotropy-robust methods (SD: −0.04; 95% CI: −0.05, −0.04 in MR-Egger and SD: −0.04;95% CI: −0.06, −0.03 per doubling in odds of genetic liability to AD for both weighted median and mode methods. The association with hippocampal volume attenuated across the pleiotropy-robust methods (SD: −0.02; 95% CI: −0.06,0.02 in the MR Egger, −0.02; 95% CI: −0.05, 0.01 in weighted mode, and SD: −0.02; 95% CI: −0.05, 0.02 per doubling in odds of genetic liability to AD in weighted median methods). In the analysis of genetic liability to AD on brain structures of participants in the UK Biobank, the magnitude of effect sizes for the MR-Egger, weighted median and mode were consistent with the IVW estimates for all brain structures (Supplementary Table 13). As expected, for some brain structures (e.g. entorhinal in the oldest age group, thalamus in the middle age group), the precision of MR-Egger was lower than the IVW due to the additional estimation of an intercept in the MR-Egger model. For putamen, caudal middle frontal, cuneus (middle age group), inferior temporal (youngest age tertile), the effect sizes of the MR-Egger, weighted median and mode were consistent with the IVW, but the 95% confidence intervals did not overlap with the null (Supplementary Table 6).

The directionality test indicated that, on average, the instruments for AD explained more variance in AD than they did in the brain structures in UK Biobank (Supplementary Table 15). When we removed the two SNPs tagging the *APOE* region from our analyses in the childhood cohort meta-analysis, the effect observed for hippocampal volume attenuated to the null, while the effect observed for the caudal anterior cingulate region remained with less precision (Supplementary Table 20). In UK Biobank analyses, the associations with regional cortical thickness and subcortical structures largely remained, but as expected, the CIs widened (Supplementary Table 21). In the MR analysis of brain structures on AD, the detrimental effect of a larger hippocampal volume on AD was consistent across weighted mode and median sensitivity analyses (Supplementary Table 22). The direction of effect was similar for the MR-Egger method but was less precisely estimated, as a result of reduced power. The directionality test showed that the hypothesised direction of the hippocampus, lateral occipital, and rostral anterior cingulate cortices on AD was true (i.e., that the instruments for these structures explain more variance in these structures than in AD risk) (Supplementary Table 22).

## Discussion

Our findings suggest that AD risk alleles have an age-dependent effect on a range of cortical and subcortical brain measures across mid- to late adulthood, but evidence for such effects in childhood, adolescence and young adulthood is weak. Our findings therefore suggest that genetic liability to AD operates through altering mechanisms and/or rates of neurodegeneration, rather than through reducing structural brain reserve. In the age-stratified analysis of UK Biobank participants, higher genetic liability to AD was associated with an age-dependent decrease in the thickness of the middle temporal, inferior temporal cortices, as well as volume of structures such as the hippocampus, accumbens and thalamus. Some effects were only apparent in the oldest participants (68-81 years), such as the decrease in the thickness of the fusiform, entorhinal and parahippocampal cortices and the volume of the amygdala. When SNPs in the *APOE* gene region were removed, effects across all structures largely remained but as expected, became less precise. In the reverse direction, we observed little evidence that the thickness and volume of cortical subcortical structures, respectively, affected the risk of AD, except for a greater hippocampal volume increasing risk.

Only weak evidence supported the association between genetic liability to AD and a smaller hippocampal volume and a lower thickness in the caudal anterior cingulate in childhood, and for the former the effect attenuates when variants in the *APOE* gene region are removed from the analysis. The associations identified with hippocampal volume are in agreement with previous PRS studies using considerably lower sample sizes and liberal p-value thresholds for SNP inclusion (increasing risk of bias due to horizontal pleiotropy). Axelrud et al^24^ and Foley et al^25^ found that genetic liability to AD was associated with lower hippocampal volume at PRS p-value thresholds for SNP inclusion at p≤0.132 and p≤0.0001, using samples with 716 and 272 participants, respectively. They concluded that the genetic effects of AD were not driven by the *APOE* gene but that the effect was due to an aggregation of smaller effects across multiple genes in the PRS. The study by Foley et al^25^ found little evidence of associations with the other structures examined (entorhinal cortex, parahippocampal gyrus, posterior cingulate gyrus, average cortical thickness and intracranial volume).

In adults, genetic liability to AD was associated with regions known to show significant atrophy early in disease progression, such as the entorhinal cortex^26–28^, the parahippocampal cortex^29^, and the hippocampus^30^. Change in hippocampal volume is an important imaging phenotype to define preclinical stages of AD, where atrophy predicts conversion from mild cognitive impairment to AD^31^. We observed a trend of a higher genetic liability to AD being associated with a smaller hippocampus in the younger participants, of ages 45-68 years. We only identified strong evidence of an effect of genetic liability on a lower hippocampal volume in the oldest age participants (68-81 years), using genetic instruments both including and excluding the *APOE* locus. A study also using the UK Biobank identified strong evidence of an effect of the AD PRS (5×10^−8^) and hippocampal subfield volumes in older individuals (63-80 years), which was driven by SNPs in the *APOE* locus^30^.

The focus of previous PRS studies with brain MRI data on the hippocampus and the neocortex can be attributed to their well-recognised role in cognition and episodic memory^32,33^. However, there are other structures that are relevant for cognition that are less well studied in relation to genetic liability to AD^34^, such as the thalamus. The medial temporal lobe connects to thalamic nuclei and the retrosplenial cortex, constituting the hippocampal-diencephalic system, whose integrity is important for normal episodic memory^35^. In our study, we found the earliest, most robust evidence of genetic liability to AD on brain structures to be in the hippocampus, caudate, accumbens, and thalamus at 60 years of age. A study investigating how the *APOE* genotype changes whole-brain large-scale structural networks in subjects with mild cognitive impairment^36^, found *APOE* ε4 carriers showed pronounced atrophy in specific regions such as the thalamus and the hippocampus, both of which had strong structural covariance association with the left caudate nucleus. A longitudinal brain imaging study^37^ examining the effects of the *APOE* ε4 genotype found evidence of differences between carriers/non-carriers in rates of amyloid-β plaque accumulation across the adult lifespan only in the caudate at age 56 years and the putamen at 63 years. *APOE* ε4 carriers showed accelerated rates of amyloid-beta deposition in the entorhinal cortex at age 68 years. We observed that the oldest participants (aged 68-81 years) with higher genetic liability to AD showed, on average, lower entorhinal thickness.

Like other studies, we also found causal effects of genetic liability to AD on larger thickness in the lateral occipital, which is consistent with two previous studies^38,39^ in healthy individuals where *APOE* ε4 carriers have a thicker occipital cortex in comparison to normal controls. The thickening of certain brain regions has been speculated to reflect brain swelling in response to glial activation in preclinical AD stages^40^.

Genetic liability to AD is hypothesised to affect brain structures through two ways: either the genetic variants influence neurodevelopment, resulting in structural differences in the brain which may increase tolerance to pathology (i.e. altering brain reserve and increase age of disease onset) or they change rates or mechanisms of neurodegeneration^3^. We observe an age-dependent decrease in the volume of structures such as the thalamus, caudate and accumbens in UK Biobank participants agreeing with the variable neurodegeneration hypothesis. Walhovd et al.^41^ examined the association between AD PRS and hippocampal volume in 1,181 cognitively healthy people with a wide age range (4–95 years). They identified an effect of a higher AD PRS on reduced hippocampal volumes in a sample of young adults, which was consistent across age groups. They did not find strong interaction effects with age, suggesting the AD PRS results in an earlier onset of brain ageing instead of accelerated ageing through variable neurodegeneration. Little evidence from our study supported the notion that brain structure alterations change the risk for AD, except for a larger hippocampal volume increasing the risk for AD.

While previous studies have examined whether genetic liability to AD is associated with specific structural brain measures, our study is the first to examine these in such large samples, using an exploratory approach from childhood to old age. Furthermore, using aggregate PRS (as previous studies have done) precludes the examination of key potential sources of bias such as horizontal pleiotropy, which we have examined in detail here. In our study, we examined regions that have not been shown to be vulnerable to AD pathology, allowing us to discover novel regions affected by genetic liability to AD, such as the caudate. The large samples of modern biobanks with neuroimaging and genetic data allowed us to recreate to the best of our ability a pseudo-longitudinal cohort. The precision of age-dependent dose effects suggest that our results are unlikely to be due to chance or other forms of bias. However, for studies such as ALSPAC, participants were selected for imaging for 1) a case-control study of psychotic experiences, 2) recall-by-genotype for schizophrenia, 3) testosterone study, making the ALSPAC sample unrepresentative of the general population. Another limitation is that different Freesurfer versions were used across cohorts. However, we allowed for this technical variation using random-effects meta-analyses. Although we applied multiple correction strategies controlling the false discovery rate, our findings were consistent across multiple cohorts.

Our study shows that genetic liability to AD is associated with age-dependent changes in brain morphology in non-clinical populations, starting as early as 60 years of age. Furthermore, we identified structures in these populations that are not shown to be affected by AD early in the course of the disease, potentially highlighting the earliest phenotypic manifestations of the disease and the optimal timing for intervention with any potential neuroprotective therapy. The lack of evidence to support an effect of brain morphology on AD suggests that genetic liability to AD affects biological pathways leading to neurodegeneration rather than neurodevelopment. Future research should aim to use a longitudinal design and integrate their findings with biological and clinical data, to prevent the incidence and progression of AD.

## Methods

### Data

#### AD GWAS

We used the largest GWAS of clinically diagnosed AD by Kunkle et al^21^. This GWAS is an extension to the original IGAP dataset by conducting a GWAS meta-analysis of non-Hispanic Whites using a larger Stage 1 discovery sample (46 datasets, n=21,982 cases, 41,944 controls), the IGAP (Alzheimer Disease Genetics Consortium (ADGC), Cohorts for Heart and Aging Research in Genomic Epidemiology Consortium (CHARGE), the European Alzheimer’s Disease Initiative (EADI), and Genetic and Environmental Risk for Alzheimer’s disease Consortium (GERAD). The analysis consisted of three stages; Stage 1 meta-analysis was followed by Stage 2, using the I-select chip (including 11,632 variants, n=18,845) and Stage 3 A (n=11,666) or Stage 3 B (N=30,511). (for SNPs not well-captured on the I-select chip). We extracted the effects of SNPs from the meta-analysis of Stage 1 and 2 and Stage 3 where available (i.e. used the effects from Stage with largest sample size), which identified 27 SNPs to be associated with AD risk.

#### Brain structure GWAS

We used GWAS of different brain structures (average thickness of the 34 cortical regions of interest, mean thickness, estimated total intracranial volume, seven subcortical volumes and total volume of white matter) conducted within different cohorts, at different ages across the life course. We included thickness rather than surface area as regional thickness has been used to differentiate between mild cognitive impairment and AD individuals with excellent accuracy, specificity, and reproducibility across independent cohorts^42^. We ran all of the GWAS described below, except for the GWAS in the ENIGMA consortium which have been previously published ^10,43–45^. Details of all GWAS conducted can be found in the Supplementary Material. GWAS for regional cortical thickness and subcortical volumes were adjusted for global cortical thickness and estimated total intracranial volume, respectively. For the peri-pubertal period, we used Generation R (a prospective population-based birth cohort from Rotterdam, the Netherlands, n=1,175, age range 8.71 to 11.99), the Adolescent Brain Cognitive Development study (ABCD) (n with imaging data= 5,022, age range 8.92 to 11.00 at baseline and IMAGEN^19^ (a multi-centre genetic neuroimaging study recruiting adolescents from secondary schools across Europe, n with imaging data=1,739, age range = 12.94 to 16.04). For early adulthood, we used The Avon Longitudinal Study of Parents and Children (ALSPAC)^46–48^, a prospective birth cohort from South West of England, n with imaging data=776, age range 18.00 to 24.5 years. For adulthood, we used the UK Biobank and stratified the sample into three equal-sized age tertiles, to examine age-specific effects. The UK Biobank is a population-based study of 503,325 participants who were recruited from across Great Britain between 2006 and 2010 (n with imaging data=9,377 per tertile, youngest age tertile = 45 to 60, middle age tertile = 60 to 68 and oldest age tertile = 68 to 81 years). Finally, we used summary data from the ENIGMA consortium ^10,12^, which includes various studies, approximately 75% are population-based and the remainder used case-control designs for various neuropsychiatric or neurodegenerative disorders (n= 33,392, age range 3.4-91.4 years). It includes the first release of UK Biobank imaging data. Table 1 summarises each of the brain structures, the number of SNPs used as instruments for each one and the accompanying F statistics. Full details of each of the cohorts, including the genotyping and neuroimaging procedures are provided in Supplementary Tables 1 and 2, respectively.

### Statistical Analyses

#### Primary analysis: Estimating the causal effect of genetic liability to AD on brain structures

##### Two sample MR

MR is a form of instrumental variable analysis to estimate the causal effect of an exposure on an outcome, using SNPs as instruments for the exposure. Two sample MR^49^ is an extension where the effects of the genetic instrument on the exposure and on the outcome are extracted from separate GWAS studies. To examine the effects of genetic liability to AD on structural brain measures, we extracted SNPs strongly associated with AD at p≤5×10^−8 21^. Where SNPs were not available, we used proxy SNPs at r^2^>0.80. SNPs were clumped using r^2^>0.001 and a physical distance for clumping of 10,000 kb. We also included rs7412 and rs423958 to tag the *APOE ε4* allele. Of the 27 SNPs identified to be strongly associated with AD, we used 23-25 SNPs as instruments for AD, the number varying according to availability within each cohort (Supplementary Table 2). We harmonized the AD and brain structure GWASs in IMAGEN, Generation R, ABCD, ALSPAC and the UK Biobank (details in Supplementary methods). We then employed univariable MR to estimate the effect of the AD SNPs on nine subcortical volumes and 34 cortical regions defined by the Desikan-Killiany atlas^50^ (as well as total volume of white matter where available) within each cohort. We used a random effects inverse-variance weighted regression analysis, which assumes no directional horizontal pleiotropy^13^. We used the F-statistic as a measure of instrument strength^51^. All effect estimates reflect standard deviation (SD) changes in the outcome per doubling of genetic liability to AD ^52^. Using the metagen function of the meta package^53^, we applied random-effects models to meta-analyse the effects of the Alzheimer’s SNPs on structural brain measures for the three peri-pubertal cohorts: IMAGEN, ABCD, and Generation R (ages 9-16 years) (Figure 1). To examine the strength of evidence of an age-related decline in the thickness and volume of cortical regions and subcortical structures, respectively, across the three age-stratified tertiles of UK Biobank, we extracted a p for difference between groups, using the meta regress command in STATA 16^54^ by classifying the model by the mean age of each tertile.

#### Secondary analyses: Estimating the causal effect of brain structures on risk of AD

##### Two-sample MR

Using the ENIGMA GWAS (i.e. the largest GWAS of brain structures)^10,12,44,45^, we extracted SNPs associated with eight subcortical volumes and the thickness of the 34 regions of interest as defined by the Desikan-Killiany atlas^50^, at genome-wide significance (5×10^−08^). SNPs were clumped using r^2^>0.001 and a physical distance for clumping of 10,000 kb. We harmonized the ENIGMA and AD GWAS (Supplementary data file). Again, we employed univariable MR to examine the causal effects of each brain structure on risk of AD using a random effects inverse variance weighted regression. All effect estimates represent an odds ratio for AD per standard deviation increase in thickness or volume.

##### Sensitivity analyses

We conducted a range of sensitivity analyses to examine for potential violation of key MR assumptions. Inverse variance weighted regression assumes no horizontal pleiotropy and provides unbiased causal effect estimates only when there is balanced or no horizontal pleiotropy. We compared estimates from inverse variance weighted to those from Egger regression^55,56^, weighted median^57^ and weighted mode^58^, which relax this assumption. Heterogeneity in the causal estimates (which can be indicative of pleiotropy) was assessed using Cochran’s Q statistic^55^. Furthermore, to exclude the possibility that the genetic instruments used to proxy for AD are instruments for brain structures and vice versa (i.e. to test that the hypothesized causal direction was correct for each SNP used), we performed a directionality (Steiger) test^59^. Where the hypothesised direction was false, we performed sensitivity analyses removing SNPs explaining more variance in the outcome than the exposure to examine change in the effects. More details on these sensitivity methods can be found in the Supplementary material. Lastly, we excluded the two SNPs in the *APOE* locus from the AD genetic instrument, to investigate whether the effects observed are driven by the variants in the *APOE* gene on chromosome 19. This study involves evaluating global patterns of effect estimates; hence, we focus on effect size and precision^60,61^. We provide adjusted p-values, controlling for the false discovery rate in the Supplementary material (Tables 4, 6 and 8).

## Supporting information

Supplementary Results

## Data Availability

Summary statistics for Alzheimers disease were obtained from the NIAGADS platform. ENIGMA MRI summary measures from genetic association analyses of estimated total intracranial volume, subcortical structures, as well as cortical thickness were requested online at http://enigma.usc.edu/research/download-enigma-gwas-results/. The ABCD Study data are openly available to qualified researchers for free. Access can be requested at https://nda.nih.gov/abcd/request-access. Requests for Generation R data should be directed toward the management team of the Generation R Study, which has a protocol of approving data requests. For access to IMAGEN data, researchers may submit a request to the IMAGEN consortium: https://imagen-europe.com/resources/imagen-project-proposal/. ALSPAC details and data descriptions are available at www.bristol.ac.uk/alspac/researchers/access where applications for individual-level data can be made (managed access). UK Biobank data are available through a procedure described at http://www.ukbiobank.ac.uk/using-the-resource/.

## Data availability

Summary statistics for AD were obtained from the NIAGADS platform. ENIGMA MRI summary measures from genetic association analyses of estimated total intracranial volume, subcortical structures, as well as cortical thickness were requested online at http://enigma.usc.edu/research/download-enigma-gwas-results/. The ABCD Study data are openly available to qualified researchers for free. Access can be requested at https://nda.nih.gov/abcd/request-access. Requests for Generation R data should be directed toward the management team of the Generation R Study, which has a protocol of approving data requests. For access to IMAGEN data, researchers may submit a request to the IMAGEN consortium: https://imagen-europe.com/resources/imagen-project-proposal/. ALSPAC details and data descriptions are available at www.bristol.ac.uk/alspac/researchers/access where applications for individual-level data can be made (managed access). UK Biobank data are available through a procedure described at http://www.ukbiobank.ac.uk/using-the-resource/.

## Code availability

Code for performing the analyses is included at https://github.com/rskl92/AD_BRAIN_BIDIRECTIONAL_MR.

## Acknowledgments

Data used in the preparation of this article were obtained from the Adolescent Brain Cognitive Development (ABCD) Study (https://abcdstudy.org), held in the NIMH Data Archive (NDA). This is a multisite, longitudinal study designed to recruit more than 10,000 children age 9-10 years and follow them over 10 years into early adulthood. The ABCD Study is supported by the National Institutes of Health and additional federal partners under award numbers U01DA041048, U01DA050989, U01DA051016, U01DA041022, U01DA051018, U01DA051037, U01DA050987, U01DA041174, U01DA041106, U01DA041117, U01DA041028, U01DA041134, U01DA050988, U01DA051039, U01DA041156, U01DA041025, U01DA041120, U01DA051038, U01DA041148, U01DA041093, U01DA041089, U24DA041123, U24DA041147. A full list of supporters is available at https://abcdstudy.org/federal-partners.html. A listing of participating sites and a complete listing of the study investigators can be found at https://abcdstudy.org/consortium_members/. ABCD consortium investigators designed and implemented the study and/or provided data but did not necessarily participate in analysis or writing of this report. This manuscript reflects the views of the authors and may not reflect the opinions or views of the NIH or ABCD consortium investigators. The ABCD data repository grows and changes over time. The ABCD data repository grows and changes over time. The ABCD data used in this report came from [NIMH Data Archive Digital Object Identifier (10.15154/1503209)].

We are extremely grateful to all the families who took part in this study, the midwives for their help in recruiting them, and the whole ALSPAC team, which includes interviewers, computer and laboratory technicians, clerical workers, research scientists, volunteers, managers, receptionists and nurses. The UK Medical Research Council and Wellcome (Grant ref: 217065/Z/19/Z) and the University of Bristol provide core support for ALSPAC. This publication is the work of RKL and ELA and NMD will serve as guarantors for the contents of this paper. A comprehensive list of grants funding is available on the ALSPAC website (http://www.bristol.ac.uk/alspac/external/documents/grant-acknowledgements.pdf). The ALSPAC-Testosterone study was funded by the National Institutes of Health, U.S.A. (R01MH085772 to TP). The ALSPAC-PE study was funded by a grant from the UK Medical Research Council (G0901885). ASD was also supported by the National Institutes of Health Research Biomedical Research Centre at the South London & Maudsley Hospital Foundation NHS Trust and the IoPPN, King’s College London. The ALSPAC SCZ-RbG study was funded by grant MR/K004360/1 from the Medical Research Council (MRC) titled: “Behavioural and neurophysiological effects of schizophrenia risk genes: a multi-locus, pathway based approach” and by the MRC Centre for Neuropsychiatric Genetics and Genomics (G0800509) and the NIHR Bristol Biomedical Research Centre. TML is supported by a Ser Cymru II Fellowship (European Regional Development Funds: CU149: “Imaging immunity in the genetic risk for Alzheimer’s disease) at the Dementia Research Institute at Cardiff University and Wellcome Trust Institutional Strategic Support Funds (513688). SD is supported by a Marie Skłodowska-Curie Actions COFUND fellowship. The MRI Pipeline development was funded by MRC Mental Health Pathfinder grant ‘Cohorts as Platforms for Mental Health research’ (MC_PC_17210).

IMAGEN is supported by the European Union-funded FP6 Integrated Project IMAGEN (Reinforcement-related behaviour in normal brain function and psychopathology) (LSHM-CT- 2007-037286), the Horizon 2020 funded ERC Advanced Grant ‘STRATIFY’ (Brain network based stratification of reinforcement-related disorders) (695313), Human Brain Project (HBP SGA 2, 785907, and HBP SGA 3, 945539), the Medical Research Council Grant ‘c-VEDA’ (Consortium on Vulnerability to Externalizing Disorders and Addictions) (MR/N000390/1), the National Institute of Health (NIH) (R01DA049238, A decentralized macro and micro gene-by-environment interaction analysis of substance use behavior and its brain biomarkers), the National Institute for Health Research (NIHR) Biomedical Research Centre at South London and Maudsley NHS Foundation Trust and King’s College London, the Bundesministeriumfür Bildung und Forschung (BMBF grants 01GS08152; 01EV0711; Forschungsnetz AERIAL 01EE1406A, 01EE1406B; Forschungsnetz IMAC-Mind 01GL1745B), the Deutsche Forschungsgemeinschaft (DFG grants SM 80/7-2, SFB 940, TRR 265, NE 1383/14-1), the Medical Research Foundation and Medical Research Council (grants MR/R00465X/1 and MR/S020306/1), the National Institutes of Health (NIH) funded ENIGMA (grants 5U54EB020403-05 and 1R56AG058854-01). Further support was provided by grants from: – the ANR (ANR-12-SAMA-0004, AAPG2019 – GeBra), the Eranet Neuron (AF12-NEUR0008-01 – WM2NA; and ANR-18-NEUR00002-01 – ADORe), the Fondation de France (00081242), the Fondation pour la Recherche Médicale (DPA20140629802), the Mission Interministérielle de Lutte-contre-les-Drogues-et-les-Conduites-Addictives (MILDECA), the Assistance-Publique-Hôpitaux-de-Paris and INSERM (interface grant), Paris Sud University IDEX 2012, the Fondation de l’Avenir (grant AP-RM-17-013), the Fédération pour la Recherche sur le Cerveau; the National Institutes of Health, Science Foundation Ireland (16/ERCD/3797), U.S.A. (Axon, Testosterone and Mental Health during Adolescence; RO1 MH085772-01A1), and by NIH Consortium grant U54 EB020403, supported by a cross-NIH alliance that funds Big Data to Knowledge Centres of Excellence.

The UK Biobank data used in this work were obtained from UK Biobank Data Application 48970. We thank UK Biobank for making the data available, and to all UK Biobank study participants, who generously donated their time to make this resource possible.

We thank the International Genomics of Alzheimer’s Project (IGAP) for providing summary results data for these analyses. The investigators within IGAP contributed to the design and implementation of IGAP and/or provided data but did not participate in analysis or writing of this report. IGAP was made possible by the generous participation of the control subjects, the patients, and their families. The i–Select chips was funded by the French National Foundation on Alzheimer’s disease and related disorders. EADI was supported by the LABEX (laboratory of excellence program investment for the future) DISTALZ grant, Inserm, Institut Pasteur de Lille, Université de Lille 2 and the Lille University Hospital. GERAD/PERADES was supported by the Medical Research Council (Grant nº 503480), Alzheimer’s Research UK (Grant n° 503176), the Wellcome Trust (Grant n° 082604/2/07/Z) and German Federal Ministry of Education and Research (BMBF): Competence Network Dementia (CND) grant n° 01GI0102, 01GI0711, 01GI0420. CHARGE was partly supported by the NIH/NIA grant R01 AG033193 and the NIA AG081220 and AGES contract N01–AG–12100, the NHLBI grant R01 HL105756, the Icelandic Heart Association, and the Erasmus Medical Center and Erasmus University. ADGC was supported by the NIH/NIA grants: U01 AG032984, U24 AG021886, U01 AG016976, and the Alzheimer’s Association grant ADGC–10–196728.

## Contributions

RKL, ELA, NMD, LD and YBS designed, conceptualised and interpreted results of the study. RKL and BX performed the statistical analyses. EC, EW, TS, TW, AW provided advice regarding structural brain measures. All authors provided valuable feedback and comments on the manuscript.

## References

1. Yang, J. et al. Voxelwise meta-analysis of gray matter anomalies in Alzheimer’s disease and mild cognitive impairment using anatomic likelihood estimation. J. Neurol. Sci. (2012). doi:10.1016/j.jns.2012.02.010

2. Bateman, R. J. et al. Clinical and Biomarker Changes in Dominantly Inherited Alzheimer’s Disease. N. Engl. J. Med. 367, 795–804 (2012).

3. Stern, Y. Cognitive reserve in ageing and Alzheimer’s disease. Lancet Neurol. 11, 1006–1012 (2012).

4. Grasby, K. L. et al. The genetic architecture of the human cerebral cortex. Science (80-.). 367, eaay6690 (2020).

5. Jansen, I. E. et al. Genome-wide meta-analysis identifies new loci and functional pathways influencing Alzheimer’s disease risk. Nat. Genet. 51, 404– 413 (2019).

6. Kunkle, B. W. et al. Genetic meta-analysis of diagnosed Alzheimer’s disease identifies new risk loci and implicates Aβ, tau, immunity and lipid processing. Nat. Genet. 51, 414–430 (2019).

7. Lambert, J.-C. Meta-Analysis of 74,046 Individuals Identifies 11 New Susceptibility Loci for Alzheimer’s Disease. Nat. Genet. 45, 1452–1458 (2013).

8. Corder, E. H. et al. Gene dose of apolipoprotein E type 4 allele and the risk of Alzheimer’s disease in late onset families. Science (80-.). 261, 921–923 (1993).

9. Gatz, M. et al. Role of genes and environments for explaining Alzheimer disease. Arch. Gen. Psychiatry 63, 168–174 (2006).

10. Grasby, K. L. et al. The genetic architecture of the human cerebral cortex. Science (80-.). 367, (2020).

11. Mormino, E. C. et al. Polygenic risk of Alzheimer disease is associated with early- and late-life processes. Neurology 87, 481–488 (2016).

12. Satizabal, C. L. et al. Genetic architecture of subcortical brain structures in 38,851 individuals. Nat. Genet. (2019). doi:10.1038/s41588-019-0511-y

13. Davey Smith, G. & Hemani, G. Mendelian randomization: genetic anchors for causal inference in epidemiological studies. Hum. Mol. Genet. 23, R89– 98 (2014).

14. Lawlor, D. A. Commentary: two-sample Mendelian randomization: opportunities and challenges. Int. J. Epidemiol. 45, 908–15 (2016).

15. Casey, B. J. et al. The Adolescent Brain Cognitive Development (ABCD) study: Imaging acquisition across 21 sites. Developmental Cognitive Neuroscience 32, 43–54 (2018).

16. Hagler, D. J. et al. Image processing and analysis methods for the Adolescent Brain Cognitive Development Study. Neuroimage 202, (2019).

17. White, T. et al. Paediatric population neuroimaging and the Generation R Study: the second wave. Eur. J. Epidemiol. (2018). doi:10.1007/s10654-017-0319-y

18. White, T. et al. Automated quality assessment of structural magnetic resonance images in children: Comparison with visual inspection and surface-based reconstruction. Hum. Brain Mapp. (2018). doi:10.1002/hbm.23911

19. Schumann, G. et al. The IMAGEN study: Reinforcement-related behaviour in normal brain function and psychopathology. Molecular Psychiatry 15, 1128–1139 (2010).

20. Sharp, T. H. et al. Population neuroimaging: generation of a comprehensive data resource within the ALSPAC pregnancy and birth cohort. Wellcome Open Res. (2020). doi:10.12688/wellcomeopenres.16060.1

21. Kunkle, B. W. et al. Genetic meta-analysis of diagnosed Alzheimer’s disease identifies new risk loci and implicates Aβ, tau, immunity and lipid processing. Nat. Genet. 51, 414–430 (2019).

22. Schöll, M. et al. PET Imaging of Tau Deposition in the Aging Human Brain. Neuron (2016). doi:10.1016/j.neuron.2016.01.028

23. Braak, H. & Braak, E. Neuropathological stageing of Alzheimer-related changes. Acta Neuropathologica 82, 239–259 (1991).

24. Axelrud, L. K. et al. Polygenic Risk Score for Alzheimer’s Disease: Implications for Memory Performance and Hippocampal Volumes in Early Life. Am. J. Psychiatry 175, 555–563 (2018).

25. Foley, S. F. et al. Multimodal Brain Imaging Reveals Structural Differences in Alzheimer’s Disease Polygenic Risk Carriers: A Study in Healthy Young Adults. Biol. Psychiatry 81, 154–161 (2017).

26. Desikan, R. S. et al. Polygenic Overlap Between C-Reactive Protein, Plasma Lipids, and Alzheimer Disease. Circulation 131, 2061–2069 (2015).

27. Killiany, R. J. et al. MRI measures of entorhinal cortex vs hippocampus in preclinical AD. Neurology 58, 1188–1196 (2002).

28. Tan, C. H. et al. Polygenic hazard score, amyloid deposition and Alzheimer’s neurodegeneration. Brain 142, 460–470 (2019).

29. Köhler, S. et al. Memory impairments associated with hippocampal versus parahippocampal-gyrus atrophy: An MR volumetry study in Alzheimer’s disease. Neuropsychologia 36, 901–914 (1998).

30. Foo, H. et al. Associations between Alzheimer’s disease polygenic risk scores and hippocampal subfieldvolumes in 17,161 UK Biobank participants. Neurobiol. Aging (2020).

31. Macdonald, K. E., Bartlett, J. W., Leung, K. K., Ourselin, S. & Barnes, J. The value of hippocampal and temporal horn volumes and rates of change in predicting future conversion to ad. Alzheimer Dis. Assoc. Disord. 27, 168–173 (2013).

32. Squire, L. R., Stark, C. E. L. & Clark, R. E. The medial temporal lobe. Annu. Rev. Neurosci. 27, 279–306 (2004).

33. Diana, R. A., Yonelinas, A. P. & Ranganath, C. Imaging recollection and familiarity in the medial temporal lobe: a three-component model. Trends Cogn. Sci. 11, 379–386 (2007).

34. Aggleton, J. P., Pralus, A., Nelson, A. J. D. & Hornberger, M. Thalamic pathology and memory loss in early Alzheimer’s disease: Moving the focus from the medial temporal lobe to Papez circuit. Brain 139, 1877–1890 (2016).

35. Opitz, B. & Friederici, A. D. Interactions of the hippocampal system and the prefrontal cortex in learning language-like rules. Neuroimage 19, 1730– 1737 (2003).

36. Novellino, F. et al. Association Between Hippocampus, Thalamus, and Caudate in Mild Cognitive Impairment APOEε4 Carriers: A Structural Covariance MRI Study. Front. Neurol. 10, (2019).

37. Mishra, S. et al. Longitudinal brain imaging in preclinical Alzheimer disease: Impact of APOE 14 genotype. Brain 141, 1828–1839 (2018).

38. Espeseth, T. et al. Accelerated age-related cortical thinning in healthy carriers of apolipoprotein E ε4. Neurobiol. Aging 29, 329–340 (2008).

39. Espeseth, T. et al. Apolipoprotein E ε4-related thickening of the cerebral cortex modulates selective attention. Neurobiol. Aging 33, 304–322 (2012).

40. Calsolaro, V. & Edison, P. Neuroinflammation in Alzheimer’s disease: Current evidence and future directions. Alzheimer’s Dement. 12, 719–732 (2016).

41. Walhovd, K. B. et al. Genetic risk for Alzheimer disease predicts hippocampal volume through the human lifespan. Neurol. Genet. 6, e506 (2020).

42. Desikan, R. S. et al. Automated MRI measures identify individuals with mild cognitive impairment and Alzheimer’s disease. Brain 132, 2048–2057 (2009).

43. Satizabal, C. L. et al. Genetic architecture of subcortical brain structures in 38,851 individuals. Nat. Genet. 51, 1624–1636 (2019).

44. Hibar, D. P. et al. Novel genetic loci associated with hippocampal volume. Nat. Commun. 8, 13624 (2017).

45. Adams, H. H. H. et al. Novel genetic loci underlying human intracranial volume identified through genome-wide association. Nat. Neurosci. 19, 1569– 1582 (2016).

46. Boyd, A. et al. Cohort profile: The ‘Children of the 90s’-The index offspring of the avon longitudinal study of parents and children. Int. J. Epidemiol. 42, 111–127 (2013).

47. Fraser, A. et al. Cohort Profile: the Avon Longitudinal Study of Parents and Children: ALSPAC mothers cohort. Int. J. Epidemiol. 42, 97–110 (2013).

48. Northstone, K. et al. The Avon Longitudinal Study of Parents and Children (ALSPAC): an update on the enrolled sample of index children in 2019 [version 1; peer review: 2 approved]. Wellcome Open Res. (2019). doi:10.12688/wellcomeopenres.15132.1

49. Lawlor, D. A. Commentary: Two-sample Mendelian randomization: Opportunities and challenges. Int. J. Epidemiol. 45, 908–915 (2016).

50. Desikan, R. S. et al. An automated labeling system for subdividing the human cerebral cortex on MRI scans into gyral based regions of interest. Neuroimage (2006). doi:10.1016/j.neuroimage.2006.01.021

51. Burgess, S. & Thompson, S. G. Avoiding bias from weak instruments in mendelian randomization studies. Int. J. Epidemiol. 40, 755–764 (2011).

52. Burgess, S. & Labrecque, J. A. Mendelian randomization with a binary exposure variable: interpretation and presentation of causal estimates. Eur. J. Epidemiol. 33, 947–952 (2018).

53. Schwarzer G. meta: An R package for meta-analysis. R News 7, 40–45 (2015).

54. Stata Press. Stata Statistical Software: Release 16. StataCorp LLC (2019).

55. Bowden, J., Davey Smith, G. & Burgess, S. Mendelian randomization with invalid instruments: Effect estimation and bias detection through Egger regression. Int. J. Epidemiol. 44, 512–525 (2015).

56. Egger, M., Davey Smith, G., Schneider, M. & Minder, C. Bias in meta-analysis detected by a simple, graphical test. BMJ 315, 629–34 (1997).

57. Bowden, J., Davey Smith, G., Haycock, P. C. & Burgess, S. Consistent Estimation in Mendelian Randomization with Some Invalid Instruments Using a Weighted Median Estimator. Genet. Epidemiol. (2016). doi:10.1002/gepi.21965

58. Hartwig, F. P., Smith, G. D. & Bowden, J. Robust inference in summary data Mendelian randomization via the zero modal pleiotropy assumption. Int. J. Epidemiol. (2017). doi:10.1093/ije/dyx102

59. Hemani, G., Tilling, K. & Davey Smith, G. Orienting the causal relationship between imprecisely measured traits using GWAS summary data. PLoS Genet. 13, (2017).

60. American Statistical Association Releases Statement on Statistical Significance and P-Values. (2016). doi:10.1080/00031305.2016.1154108#.Vt2XIOaE2MN

61. Sterne, J. A. C. & Davey Smith, G. Sifting the evidence—what’s wrong with significance tests? BMJ 322, 226 (2001).

