## Supplementary Methods for "The bidirectional causal effects of brain morphology across the life course and risk of Alzheimer’s disease: A cross-cohort comparison and Mendelian randomization meta-analysis"

### Cohort description

#### ABCD

The ABCD study ([http://abcdstudy.org](http://abcdstudy.org/)) consists of 11,875 participants of ages 9-10 years old at baseline. The aim of this study was to examine the effect of brain structure and function on developmental trajectories and addiction^1^.

#### IMAGEN

The IMAGEN study is a European multi-centre genetic neuroimaging study recruiting adolescents from secondary schools in London, Nottingham, Dublin, Paris, Berlin, Hamburg, Mannheim and Dresden. To ascertain a diverse sample with respect to socioeconomic status, emotional and cognitive development, private, state-funded and special schools were equally targeted. Participation in the study involves visits to the study centre and home assessments^2^.

#### Generation R

The Generation R study is a prospective population-based birth cohort from Rotterdam, the Netherlands. The aim of the Generation R Study is to identify genetic and environmental determinants that affect maternal and child development^3^. Study protocols were approved by the Medical Ethics Committee of the Erasmus Medical Center. All participants and their parents provided assent and informed consent, respectively.

#### ALSPAC

The Avon Longitudinal Study of Parents and Children (ALSPAC) is a prospective birth cohort which recruited pregnant women with expected delivery dates between April 1991 and December 1992 from Bristol UK^4–6^. The initial number of pregnancies enrolled is 14,541 and of these initial pregnancies, there was a total of 14,676 foetuses, resulting in 14,062 live births and 13,988 children who were alive at 1 year of age. When the oldest children were approximately 7 years of age, an attempt was made to bolster the initial sample with eligible cases who had failed to join the study originally. As a result, when considering variables collected from the age of seven onwards (and potentially abstracted from obstetric notes) there are data available for more than the 14,541 pregnancies mentioned above. The total sample size for analyses using any data collected after the age of seven is therefore 15,454 pregnancies, resulting in 15,589 foetuses. Of these 14,901 were alive at 1 year of age. Between the ages of 18 to 21 years, a subset of ALSPAC offspring were invited to participate in three different neuroimaging studies: the ALSPAC-Testosterone study, the ALSPAC-Psychotic Experiences (PE) study, and the ALSPAC- Schizophrenia Recall by Genotype (SCZ-RbG) study. In total, MRI data was acquired for 958 participants: 513 in the Testosterone study, 248 in the Psychotic Experiences study and 197 in the ALSPAC-Schizophrenia Recall by Genotype study^7^. Please note that the study website contains details of all the data that is available through a fully searchable data dictionary and variable search tool (<http://www.bristol.ac.uk/alspac/researchers/our-data/>). Ethical approval for the study was obtained from the ALSPAC Ethics and Law Committee and the Local Research Ethics Committees. Informed consent for the use of data collected via questionnaires and clinics was obtained from participants following the recommendations of the ALSPAC Ethics and Law Committee at the time.

#### UK Biobank

UK Biobank is a population-based study of 503,325 participants who were initially recruited from across Great Britain between 2006 and 2010, aged 40–69 years ([http://www.ukbiobank.ac.uk](http://www.ukbiobank.ac.uk/)). UK Biobank received ethical approval from the research ethics committee (REC reference 11/NW/0382). The present analyses were conducted under UK Biobank application number 48970.

| Table 1. Cohort image acquisition and processing | | | | | |
| --- | --- | --- | --- | --- | --- |
| **Study sample** | **MRI-scanner** | **Software** | **Acquisition** | **Reference (PMID)** | **Quality control** |
| The Adolescent Brain and Cognitive Development study (ABCD) | The ABCD imaging protocol is harmonized for three 3T scanner platforms (Siemens Prisma, General Electric (GE) 750 and Philips) | Freesurfer v6.0.0 | **Siemens:** matrix=256x256, slices=176, FOV=256x256, % FOV phase=100%,Resolution (mm)=1.0x1.0x1.0, TR (ms)= 2000, TE (ms)=2.88, TI (ms)=1060, Flip Angle (deg)=8, Parallel Imaging=2x, MultiBand Acceleration=Off, Phase partial fourir=Off, Diffusion directions=N/A, b-values=N/A, Acquisition time=7:12  **Philips:** matrix=256x256, slices=225, FOV=256x240,% FOV phase=93.75%,Resolution (mm)=1.0x1.0x1.0, TR (ms)=6.31, TE (ms)=2.9, TI (ms)=1060, Flip Angle (deg)=8, Parallel Imaging=1.5x2.2, MultiBand Acceleration=Off, Phase partial fourir=Off, Diffusion directions=N/A, b-values=N/A, Acquisition time=5:38;  **General Electric:** matrix=256x256, slices=208,FOV=256x256, % FOV phase=100%,Resolution (mm)=1.0x1.0x1.0, TR (ms)= 2500, TE (ms)=2, TI (ms)=1060, Flip Angle (deg)=8, Parallel Imaging=2x, MultiBand Acceleration=Off, Phase partial fourir=Off, Diffusion directions=N/A, b-values=N/A, Acquisition time=6:09 | 31415884 | Removal of measures based on result of visual inspection |
| Avon Longitudinal Study of Parents and Children (ALSPAC) | 3 Tesla General Electric HDx (GE Medical Systems) using an 8-channel head coil | Freesurfer v6.0.0 | During each structural imaging session coronal T 1 scans were collected. Imaging parameters were as follows: 3D fast spoiled gradient echo (FSPGR) with 168–182 oBetalique-axial AC-Precuneusslices, 1 mm isotropic resolution; flip angle = 20°; repetition time (TR) = 7.9 ms; echo time (TE) = 3.0 ms; inverse time (TI) = 450 ms; 1mm × 1mm x 1mm voxel size; slice thickness 1 mm; FOV (field of view) 256 × 192 mm matrix. T 1- weighted scans took approximately 7.15 minutes each. | 33043145 | Removal of measures based on result of visual inspection |
| Generation R | 3-Tesla MRI system (MR-750W, General Electric, Milwaukee, WI, US) using an eight-channel, receive-only head coil | Freesurfer v6.0.0 | High-resolution, T1-weighted structural MRI data were acquired using a coronal inversion recovery fast spoiled gradient recalled sequence with the following parameters: GE option BRAVO, TR = 8.77 ms, TE = 3.4 ms, TI = 600 ms, flip angle = 10°, matrix size = 220 × 220, field of view = 220 mm × 220 mm, slice thickness = 1 mm, number of slices = 230, ARC acceleration factor = 2. | 29064008 | Removal of measures based on result of visual inspection |
| IMAGEN | 3.0 T Philips Medical Systems Achieva;  3.0 T Brucker;  3.0 T Siemens TrioTim;  3.0 T Siemens Verio;  3.0 T Brucker/GE Medical Systems Signa Excite;  3.0 T GE Medical Systems Signa HDx | Freesurfer v5.3.0 | Magnetic resonance imaging data were acquired at 8 European centers, using a standardised 3 Tesla, T1-weighted gradient echo protocol (voxel size=1.1 mm isotropic) based on that from the ADNI initiative (http://adni.loni.usc.edu/methods/documents/mri-protocols/) | 21102431 | No visual inspection of MRI images had been performed. We removed outliers 3 standard deviations above/below mean of respective measures. |
| UK Biobank | Siemens Skyra 3T running VD13A SP4, with a standard Siemens 32-channel RF receive head coil | Freesrfer v5.3.0 | Voxel matrix: 1.0x1.0x1.0 mm - 208x256x256. 3D MPRAGE, TI/TR=880/2000 ms, sagittal orientation, in-plane acceleration factor=2 | 27643430, 29079522 | No visual inspection of MRI images had been performed. We removed outliers 3 standard deviations above/below mean of respective measures. |

### Genetic data

ALSPAC, IMAGEN, Generation R, and ABCD participants were genotyped using the Illumina HumanHap550, Illumina 610 and Illumina 660K, Illumina 670, and the Affymetrix NIDA SmokeScreen Array chips, respectively. For UK Biobank, the initial 50,000 participants were genotyped on Affymetrix UK BiLEVE Axiom array and the remaining 450,000 participants were genotyped using the Affymetrix UK Biobank Axiom® array and quad chip genotyping platforms. Details on quality control and imputation panels have been published elsewhere^8^. For ABCD, saliva samples were collected at baseline and sent to Rutgers University Cell and DNA Repository for storage and DNA isolation and genotyping was conducted using the Smokescreen array^9^. The initial dataset provided by the ABCD (ABCD_release_2.0.1_r1) included 517,724 genetic variants chr1-23,25-26. For ABCD, we performed quality control using plinkQC and imputation of the genetic data using the Michigan imputation server. We identified individuals of European ancestry by combining the genotypes of ABCD with genotypes of 1000 genomes phase 3, consisting of individuals from known ethnicities. Principal component analysis using this genotype panel can be used to identify population structure down to the level of 1000 genomes (i.e. large-scale continental ancestry). To identify these individuals, we used check_ancestry implemented in PLINK QC. It uses principal components 1 and 2 to find the centre of the European reference samples. We performed PCA analysis on the pruned ABCD dataset (Number of variants=152,094). All study samples whose euclidean distance from the centre falls outside a specified radius are considered non-European. We performed individual/sample-level quality control, as well as marker quality control. For each sample, the homozygosity rates across all X-chromosomal genetic variants were computed and compared with expected rates (females, X homozygosity<0.2); males, X chrom homozygosity>0.8). Samples with discordant sex information that is not accounted for were removed from the study. Outlying missing genotype and/or heterozygosity rates aids in detecting samples with poor DNA quality and/or concentration that should be removed from the study. We excluded individuals based a missing genotype rate of 3%, and individuals whose heterozygosity rate was 3 standard deviations above or below mean heterozygosity rate. For the marker-level quality control, we filtered genetic variants based on a Hardy Weinberg equilibrium exact test p value of 5e-07, a call rate of 95% and a minor allele frequency of 5%. In total, we retained 9,907 individuals (5,300 were of European ancestry) and 377,164 genetic variants. We performed imputation using the Michigan Imputation Server using hrc.r1.1.2016 reference panel, Eagle v2.3 phasing and multi-ethnic imputation process ^10^. Code and further details can be found here <https://github.com/rskl92/ABCD_QC_genetic_data>.

| Table 2. Information on genotyping and quality control. | | | | | | | | |
| --- | --- | --- | --- | --- | --- | --- | --- | --- |
| **Data type** | **Cohort/**  **source** | **HWE** | **MAF** | **Call Rate** | **Association** | **Imputation** | **Reference population** | **Genotype Platform** |
| Individual-level data | ABCD | 5.00E-07 | 0.01 | 0.95 | Eagle2 | minimac4 | Haplotypes Reference Consortium (HRC) | Smokescreen array |
|  | ALSPAC | 5.00E-07 | 0.01 | 0.95 | ShapeIT2 | MACH 1.0.16 Markov Chain Haplotyping | 1000 genomes phase 1 version 3 (release date 21/05/2011) | Illumina HumanHap550 quad |
|  | Generation R | 1.00E-07 | 0.001 | 0.9 | mach | minimac? | 1000 Genomes (phase 3; March 2012) | Illumina 610 and 660 K |
|  | IMAGEN | 1.00E-06 | 0.01 | 0.95 | mach2qtl(1.1.2) | minimac (release 2012-05-29) | 1000 Genomes (phase 1 version 3; Nov 2010) | Illumina 610-Quad and Illumina 660W-Quad |
|  | UK Biobank | 1.00E-06 | >3% for info>0.3; info>0.6 for MAF 1-3%, info>0.8 for MAF 0.5-1%;info>0.9 for MAF 0.1-0.5% | 0.95 | SHAPEIT2 | impute2 | Haplotypes Reference Consortium (HRC) and UK10K haplotype | UK Biobank Axiom Array |
| Summary-level data for subcortical structures | ENIGMA consortium;  Satizabal, Hibar, and Adams | See respective publication ^11–13^ | See respective publication ^11–13^ | See respective publication ^11–13^ | See respective publication ^11–13^ | See respective publication ^11–13^ | See respective publication ^11–13^ | See respective publication ^11–13^ |
| Summary-level data for cortical measures | ENIGMA consortium; Grasby et al | See respective publication ^14^ | See respective publication ^14^ | See respective publication ^14^ | See respective publication ^14^ | See respective publication ^14^ | See respective publication ^14^ | See respective publication ^14^ |

### Genome-wide association studies

#### Alzheimer’s GWAS

International Genomics of Alzheimer's Project (IGAP) is a large three-stage study based upon genome-wide association studies (GWAS) on individuals of European ancestry. In stage 1, IGAP used genotyped and imputed data on 11,480,632 single nucleotide polymorphisms (SNPs) to meta-analyse GWAS datasets consisting of 21,982 Alzheimer’s disease cases and 41,944 cognitively normal controls from four consortia: The Alzheimer Disease Genetics Consortium (ADGC); The European Alzheimer's disease Initiative (EADI); The Cohorts for Heart and Aging Research in Genomic Epidemiology Consortium (CHARGE); and The Genetic and Environmental Risk in AD Consortium Genetic and Environmental Risk in AD/Defining Genetic, Polygenic and Environmental Risk for Alzheimer’s Disease Consortium (GERAD/PERADES). In stage 2, 11,632 SNPs were genotyped and tested for association in an independent set of 8,362 Alzheimer's disease cases and 10,483 controls. Meta-analysis of variants selected for analysis in stage 3A (n = 11,666) or stage 3B (n = 30,511) samples brought the final sample to 35,274 clinical and autopsy-documented Alzheimer’s disease cases and 59,163 controls.

#### Individual-level brain GWAS

We retained only individuals of European ancestry from all cohorts used in our analysis. In ALSPAC, IMAGEN, Generation R, and UK Biobank we included unrelated individuals.

We estimated the effects of the Alzheimer’s disease genetic variants on each standardised brain structure in individuals of European ancestry, using the lm function in RStudio version 3.3.1, for ALSPAC, IMAGEN, Generation R, and UK Biobank. The models for all cohorts were adjusted for age, sex, and ancestry-derived principal components. For ABCD, we used BOLT-LMM version 2.3 (linear mixed model (LMM)) software. BOLT-LMM applies an LMM to examine the association between genetic variants and phenotypes, whilst accounting for population stratification and cryptic relatedness^15^ . For ALSPAC, IMAGEN, and Generation R, we adjusted for five genetic principal components. For ABCD and UK Biobank, we adjusted for ten genetically informative principal components.

#### Summary level brain GWAS

Summary statistics of cortical thickness and subcortical volumes were obtained from the ENIGMA consortium^11–13^. The GWAS by Satizabal et al^11^ were based on brain MRI scans and genome wide genotype data of up to 37,741 individuals from ENIGMA, CHARGE, and UK Biobank. GWAS for hippocampal volume^12^ and estimated total intracranial volume^13^ were based on the brain MRI images of 33,535 and 37,345 participants, respectively, from the ENIGMA and CHARGE consortia. The meta-analyses of global and regional thickness of 34 cortical regions consisted results from 33,992 participants from ENIGMA and UK Biobank^14^. All cortical regions and subcortical structures were mapped to the Desikan-Killiany atlas^16^. All participants in these studies provided written informed consent and sites involved obtained approval from local research ethics committees or Institutional Review Boards.

### Two-sample Mendelian randomization

Harmonization of exposure and outcome GWAS data
Only biallelic single nucleotide polymorphisms (SNPs) were included as instruments (insertions and deletions were removed). SNPs for each Alzheimer’s SNP was identified in the brain GWAS. Proxies were identified for any SNPs not found (r^2^>0.8 using 1000 genomes as a reference). Proxies may differ between datasets. In a two-sample MR analysis, the effect of a SNP on exposure and an outcome must be harmonised relative to the same allele. SNPs for the exposure were coded such that the effect allele was always the ‘increasing allele’ (e.g. allele increasing Alzheimer’s disease when examining genetic liability for Alzheimer’s disease on brain structures or the allele increasing brain structure in the reverse direction of the bidirectional analysis), and the alleles were harmonized so that the effect on the outcome corresponded to the same allele as the exposure.

Sensitivity analyses
A range of sensitivity analyses were conducted to check for violation of the key MR assumptions and check the robustness of the causal effect estimates:
(1) The inverse variance weighted (IVW) method assumes no horizontal pleiotropy (i.e. it assumes there are no causal paths from the SNPs to brain structures that do not go through Alzheimer’s disease)^17^. It also assumes the gene-exposure association estimates are measured with no measurement error (NOME assumption)^17^. Thus, we compared effect estimates from the IVW regressions to those obtained with MR-Egger regression models, as the use of many alleles in MR analyses increases the potential for pleiotropic effects due to aggregation of invalid genetic instruments^18^. MR-Egger assumes NOME but relaxes the assumption that the effects of genetic variants on the outcome operate entirely via the exposure (i.e. no horizontal pleiotropy), by not constraining the intercept term to zero in the weighted regression described above. In this instance, the intercept parameter estimates the overall pleiotropic effect of the SNPs on the outcome, with a non-zero intercept providing evidence for bias due to pleiotropy. The beta coefficient (or slope) of MR-Egger provides a causal estimate of the exposure on the outcome, accounting for this level of pleiotropy and assuming that the pleiotropic effect of SNPs on the outcome is not correlated with the instrument strength.
(2) We compared the results from IVW and MR-Egger regression to those obtained with the weighted median method^19^, which provides a consistent estimate of causal effect if at least 50% of the genetic variants are valid instrumental variables (i.e. robustly associated with the exposure, not associated with confounding factors and only associated with the outcome via the exposure of interest). The weighted mode method assumes that the plurality of genetic variants are valid instrumental variables^20^.(4) The presence of excessive between-SNP heterogeneity in an MR analysis may indicate that some of the genetic variants are pleiotropic. Thus, we assessed heterogeneity (i.e. variability in estimates from different genetic variants) using Cochran’s Q statistic^17^.

(3) In MR, it is assumed that the genetic instruments influence the exposure first and then the outcome, through the exposure. However, it is possible that the SNPs used to instrument structural brain measures may have a direct effect on AD risk, which then goes on to influence structural brain measures. To test that the hypothesised causal direction was correct for each SNP, we performed directionality tests which investigate whether the SNP explains more variance in the exposure than it does in the outcome (which should be true if the hypothesised causal direction from exposure to outcome is correct)^21^.

**References:**

1.	Casey, B. J. *et al.* The Adolescent Brain Cognitive Development (ABCD) study: Imaging acquisition across 21 sites. *Developmental Cognitive Neuroscience* **32**, 43–54 (2018).

2.	Schumann, G. *et al.* The IMAGEN study: Reinforcement-related behaviour in normal brain function and psychopathology. *Molecular Psychiatry* **15**, 1128–1139 (2010).

3.	White, T. *et al.* Pediatric population-based neuroimaging and the Generation R Study: The intersection of developmental neuroscience and epidemiology. *Eur. J. Epidemiol.* **28**, 99–111 (2013).

4.	Fraser, A. *et al.* Cohort Profile: the Avon Longitudinal Study of Parents and Children: ALSPAC mothers cohort. *Int. J. Epidemiol.* **42**, 97–110 (2013).

5.	Boyd, A. *et al.* Cohort Profile: the ’children of the 90s’--the index offspring of the Avon Longitudinal Study of Parents and Children. *Int. J. Epidemiol.* **42**, 111–127 (2013).

6.	Northstone, K. *et al.* The Avon Longitudinal Study of Parents and Children (ALSPAC): an update on the enrolled sample of index children in 2019 [version 1; peer review: 2 approved]. *Wellcome Open Res.* (2019). doi:10.12688/wellcomeopenres.15132.1

7.	Sharp, T. H. *et al.* Population neuroimaging: generation of a comprehensive data resource within the ALSPAC pregnancy and birth cohort. *Wellcome Open Res.* (2020). doi:10.12688/wellcomeopenres.16060.1

8.	Bycroft, C. *et al.* Genome-wide genetic data on ~500,000 UK Biobank participants. *doi.org* 166298 (2017). doi:10.1101/166298

9.	Baurley, J. W., Edlund, C. K., Pardamean, C. I., Conti, D. V. & Bergen, A. W. Smokescreen: A targeted genotyping array for addiction research. *BMC Genomics* (2016). doi:10.1186/s12864-016-2495-7

10.	Das, S. *et al.* Next-generation genotype imputation service and methods. *Nat. Genet.* **48**, 1284–1287 (2016).

11.	Satizabal, C. L. *et al.* Genetic architecture of subcortical brain structures in 38,851 individuals. *Nat. Genet.* (2019). doi:10.1038/s41588-019-0511-y

12.	Hibar, D. P. *et al.* Novel genetic loci associated with hippocampal volume. *Nat. Commun.* **8**, 13624 (2017).

13.	Adams, H. H. H. *et al.* Novel genetic loci underlying human intracranial volume identified through genome-wide association. *Nat. Neurosci.* **19**, 1569–1582 (2016).

14.	Grasby, K. L. *et al.* The genetic architecture of the human cerebral cortex. *Science (80-. ).* **367**, eaay6690 (2020).

15.	Loh, P. R., Kichaev, G., Gazal, S., Schoech, A. P. & Price, A. L. Mixed-model association for biobank-scale datasets. *Nat. Genet.* **50**, 906–908 (2018).

16.	Desikan, R. S. *et al.* An automated labeling system for subdividing the human cerebral cortex on MRI scans into gyral based regions of interest. *Neuroimage* **31**, 968–980 (2006).

17.	Bowden, J., Davey Smith, G. & Burgess, S. Mendelian randomization with invalid instruments: Effect estimation and bias detection through Egger regression. *Int. J. Epidemiol.* **44**, 512–525 (2015).

18.	Haycock, P. C. *et al.* Best (but oft-forgotten) practices: The design, analysis, and interpretation of Mendelian randomization studies. *American Journal of Clinical Nutrition* **103**, 965–978 (2016).

19.	Bowden, J., Davey Smith, G., Haycock, P. C. & Burgess, S. Consistent Estimation in Mendelian Randomization with Some Invalid Instruments Using a Weighted Median Estimator. *Genet. Epidemiol.* (2016). doi:10.1002/gepi.21965

20.	Hartwig, F. P., Smith, G. D. & Bowden, J. Robust inference in summary data Mendelian randomization via the zero modal pleiotropy assumption. *Int. J. Epidemiol.* (2017). doi:10.1093/ije/dyx102

21.	Hemani, G., Tilling, K. & Davey Smith, G. Orienting the causal relationship between imprecisely measured traits using GWAS summary data. *PLoS Genet.* **13**, (2017).
